# Blood pressure variability, central autonomic network dysfunction and cerebral small vessel disease in APOE4 carriers

**DOI:** 10.1101/2023.12.13.23299556

**Authors:** Trevor Lohman, Isabel Sible, Arunima Kapoor, Allison C Engstrom, John Paul Alitin, Aimee Gaubert, Kathleen E Rodgers, David Bradford, Mara Mather, S. Duke Han, Julian F. Thayer, Daniel A Nation

**Affiliations:** University of Southern California Leonard Davis School of Gerontology, Los Angeles, CA, USA; Department of Psychology, University of Southern California, Los Angeles, CA, USA; Department of Psychological Science, University of California, Irvine, Irvine, CA, USA; Center for Innovations in Brain Science, Department of Pharmacology, University of Arizona, Tucson, AZ, USA; University of Southern California Keck School of Medicine, Los Angeles, CA, USA

**Keywords:** blood pressure variability, *APOE4*, small vessel disease, central autonomic network

## Abstract

**Background:** Increased blood pressure variability (BPV) is a risk factor for cerebral small vessel disease (CSVD) and neurodegeneration, independent of age and average blood pressure, particularly in apolipoprotein E4 (*APOE4*) carriers. However, it remains uncertain whether BPV elevation is a cause or a consequence of vascular brain injury, or to what degree injury to the central autonomic network (CAN) may contribute to BPV-associated risk in *APOE4* carriers.

**Methods:** Independently living older adults (n=70) with no history of stroke or dementia were recruited from the community and underwent 5 minutes of resting beat-to-beat blood pressure monitoring, genetic testing, and brain MRI. Resting BPV, *APOE* genotype, CSVD burden on brain MRI, and resting state CAN connectivity by fMRI were analyzed. Causal mediation and moderation analysis evaluated BPV and CAN effects on CSVD in *APOE4* carriers (n=37) and non-carriers (n=33).

**Results:** Higher BPV was associated with the presence and extent of CSVD in *APOE4* carriers, but not non-carriers, independent of CAN connectivity (*B*= 18.92, *P*= .02), and CAN connectivity did not mediate the relationship between BPV and CSVD. In *APOE4* carriers, CAN connectivity moderated the relationship between BPV and CSVD, whereby BPV effects on CSVD were greater in those with lower CAN connectivity (*B*= 36.43, *P*= .02).

**Conclusions:** Older *APOE4* carriers with higher beat-to-beat BPV exhibit more extensive CSVD, independent of average blood pressure, and the strength of CAN connectivity does not mediate these effects. Findings suggest increased BPV is more likely a cause, not a consequence, of CSVD. BPV is more strongly associated with CSVD in *APOE4* carriers with lower rsCAN connectivity, suggesting CAN dysfunction and BPV elevation may have synergistic effects on CSVD. Further studies are warranted to understand the interplay between BPV and CAN function in *APOE4* carriers.

## Introduction

Increased blood pressure variability (BPV) has emerged as a risk factor for stroke (*1*), cerebral small vessel disease (CSVD) (*2, 3*), neurodegenerative disease (*4*) and dementia (*5*), independent of average blood pressure. A growing number of studies examining the adverse effects of BPV on both cerebrovascular and neurodegenerative diseases have highlighted the importance of the apolipoprotein ε4 allele (*APOE4*), the most important genetic risk factor for dementia due to neurodegeneration (*6*) and cerebrovascular disease (*7*). The *APOE4*- associated risk for dementia is thought to be due to pleiotropic effects on cerebral amyloidosis (*8*), CSVD (*9*), medial temporal blood-brain barrier disruption (*10, 11*), medial temporal atrophy (*12*), and resulting cognitive decline (*13*). Although most studies of BPV have not examined the role of *APOE4*, several reports indicate BPV-associated risk is predominantly observed in *APOE4* carriers (ε3/ε4 or ε4/ε4), including BPV-related risk for cerebral amyloidosis (*4*), CSF and plasma phosphorylated tau changes (*4, 14*), medial temporal tau accumulation (*15*) and medial temporal atrophy (*16*). To our knowledge, no studies have examined whether BPV-related CSVD changes are more prominent in *APOE4* carriers, despite the increased risk for CSVD associated with *APOE4* (*7, 17*).

There are also open questions regarding the causal directionality of BPV-associated risk for cerebrovascular disease. Many have hypothesized that BPV elevation is a modifiable risk factor that is causally linked to vascular brain injury since increased BPV precedes and predicts future longitudinal decline in medial temporal cerebral blood flow (*18*), accelerated neurodegeneration (*4, 15, 16*) and decline in cognitive ability (*19, 20*). However, causal directionality has been questioned by some (*21*) since BPV is partly determined by the baroreflex (*22*), and both neurodegeneration and white matter damage due to CSVD may be associated with autonomic dysfunction (*23*). Numerous functional brain MRI studies have identified a central autonomic network (CAN) involved in control of blood pressure (*24*), heart rate (*25*), heart rate variability (*26*) and baroreflex sensitivity (*27*). The CAN includes grey matter regions susceptible to vascular and neurodegenerative disease in *APOE4* carriers, particularly the hippocampus (*10, 28*), amygdala (*28, 29*) and insular cortices (*28, 29*). Reverse causation is plausible if neurodegeneration and vascular changes in CAN regions or their connecting white matter tracts leads to elevated BPV (*30–33*). There is also the possibility of a synergistic effect or vicious cycle of BPV elevation causing vascular brain injury that in turn disrupts the CAN and drives further increases in BPV. In support of this hypothesis, at least one study observed that both autonomic dysfunction and BPV elevation together predict dementia (*34*). To our knowledge, no studies to date have examined BPV and CAN connectivity in relation to CSVD in older *APOE4* carriers.

The present study sought to evaluate the causal directionality of associations among BPV, resting state central autonomic network (rsCAN) functional connectivity and CSVD in older *APOE4* carriers.

## Methods

### Participants

Participants were recruited from Los Angeles County and Orange County communities, and all procedures were conducted as part of the Vascular Senescence and Cognition (VaSC) Study at the University of Southern California (USC) and University of California Irvine (UCI).

Older adults aged 55 to 89 years who were living independently were included. Exclusion criteria were history of clinical stroke, dementia, major neurological or psychiatric disorder or medications impairing the central nervous system, current organ failure or other uncontrolled systemic illness, or contraindication for brain MRI. Study inclusions and exclusions were verified by a structured clinical health interview and review of current medications with the participant and, when available, an informed study partner. Seated systolic and diastolic blood pressure, height and weight were also recorded. This study was approved by the USC and UCI Institutional Review Boards, and all participants gave informed consent.

## Measures

### Blood Pressure Assessment

Beat-to-beat blood pressure measurements were obtained continuously during resting state fMRI scan with participants in the supine position using an MRI compatible device (Biopac®). Data were processed with a custom pipeline scripted in AcqKnowledge® that excluded outliers and instances of noise (e.g., signal dropout due to sensor interference), as described elsewhere (*35*). Systolic blood pressure (SBP) readings were then averaged over 5 minutes of recording, and intraindividual BPV was calculated as variability independent of mean (VIM), a commonly used index of BPV that is uncorrelated with average BP (*36–38*). VIM was calculated as standard deviation/mean^x^, where x was derived from a non-linear fitting of BP standard deviation (SD) against average BP using the nls package in R, as described elsewhere (*38*). Systolic blood pressure standard deviation (BP SD) and coefficient of variation (CV = (SD/mean) * 100) were also calculated for analysis.

### APOE Genotyping

Fasted blood samples were obtained by venipuncture and used to determine participant *APOE* genotype. Genomic DNA was extracted using the PureLink Genomic DNA Mini Kit (Thermo). The isolated DNA concentration was determined using a NanoDrop One (Thermo). DNA was then stored atL−L80 °C for long-term storage. Isolated DNA was first diluted to a concentration of 10 mg/μL. PCR reactions were performed in a final volume of 25 μL containing 25 ng DNA, 0.5 μM of both forward and reverse primers (forward: ACGGCTGTCCAAGGAGCTG; reverse: CCCCGGCCTGGTACACTG), and 1×LSYBR Green Master Mix (Qiagen) diluted in H2O. For the amplification, a T100 Thermal Cycler (BioRad) was used with the following settings: 95 °C for 10 min; 32 cycles of 94 °C for 20 s, 64 °C for 20 s, and 72 °C for 40 s; followed by 72 °C for 3 min. 15 μL of the DNA PCR product was digested with Hhal-fast enzyme at 37 °C for 15 min. The digested PRC product was added to a 3% agarose gel in 1×Lborax buffer for gel electrophoresis. The gel was run at 175 V for 25 min and visualized on ChemiDoc (BioRad) with a GelRed 10,000×Lgel dye. *APOE4* carrier status was defined as *APOE4* carriers (at least one copy of the ε4 allele) or *APOE4* non-carriers (no copies of the ε4 allele), as previously described (*39*).

### Vascular Risk Factors (VRF)

VRF were evaluated through physical exam, clinical blood tests and interviews with the participant and informant, and included history of cardiovascular disease (e.g., heart failure, angina, stent placement, coronary artery bypass graft, intermittent claudication), hypertension, hyperlipidemia, type 2 diabetes, atrial fibrillation, and transient ischemic attack (TIA), and total VRF burden was defined by the sum of these risk factors as described elsewhere (*37, 40*).

## MR Imaging Procedures

All participants underwent brain MRI scans conducted on a 3T Siemens Prisma scanner with 20-channel head coil. High-resolution 3D T1-weighted anatomical (Scan parameters: TR = 2300 ms; TE = 2.98 ms; TI = 900 ms; flip angle = 9 deg; FOV = 256 mm; resolution = 1.0 × 1.0 × 1.2 mm^3^; Scan time = 9 min) images were acquired, using 3- dimensional magnetization-prepared rapid gradient-echo (MPRAGE) sequences. Resting state fMRI scans comprised 140 contiguous echo-planar imaging (EPI) functional volumes (TR = 3,000 ms, TE = 30 ms, FA = 80°, 3.3 × 3.3 × 3.3 mm voxels, matrix = 64 × 64, FoV = 212 mm, 48 slices). T2-weighted scan parameters: TR = 10,000 ms; TE = 88.0 ms; flip angle = 120 deg; FOV = 210 mm; resolution = 0.8 × 0.8 × 3.5 mm^3^; Echo spacing = 9.8 ms; Echo trains per slice = 11; Scan time = 2 min).

### Cerebral Small Vessel Disease and White Matter Lesion (WML) Volume Quantification

The following sequences were examined for the current analysis: 3D T1-weighted anatomical scan for qualitative assessment of brain structures and abnormalities, T2-weighted scan for identification of enlarged perivascular spaces, fluid-attenuated inversion recovery (T2- FLAIR) for the evaluation of white matter hyperintensities and differentiation of lacunes and perivascular spaces, and T2∗-weighted imaging for identification of cerebral microbleeds (*41–43*). MRI markers were identified in accordance with established neuroimaging standards for CSVD (*44*) and scored by a blinded rater (AK) trained by a board-certified neuroradiologist, as previously reported (*45, 46*). To determine total MRI CSVD burden, all imaging markers were combined using a total CSVD burden score as described elsewhere (*47*), which ranges from 0 to 4 and includes presence of white matter hyperintensities (1 point for Fazekas score 2-3), lacunes (1 point for ≥1), microbleeds (1 point for ≥1) and perivascular spaces (1 point for moderate to severe basal ganglia perivascular spaces). Total CSVD burden was dichotomized into CSVD feature absence (CSVD burden total score 0) and presence (CSVD burden total score 1–4).

White matter lesions were segmented with the lesion growth algorithm implemented in the LST toolbox version 3.0.0 (www.statistical-modelling.de/lst.html) for SPM12 (*48*). Initial threshold was set at .2 and visual inspection was conducted to determine optimal threshold for each individual; manual quality control check ensured no gross over- or under-estimation.

### Selection of Central Autonomic Network (CAN) Regions of Interest

A CAN quantification method including 12 regions of interest (ROIs) associated with autonomic function was applied, as previously reported (*28*). This network was derived from a meta-analysis of 43 studies accounting for CAN heterogeneity in the literature. The included CAN ROIs have previously been implicated in autonomic regulation in both task-based and resting state fMRI studies (*26*). The 12 included ROIs are displayed in Figure 1 and are defined by 5mm spheres around the MNI coordinates listed in **Table 1** using the CONN Functional Toolbox.

**Figure 1:**
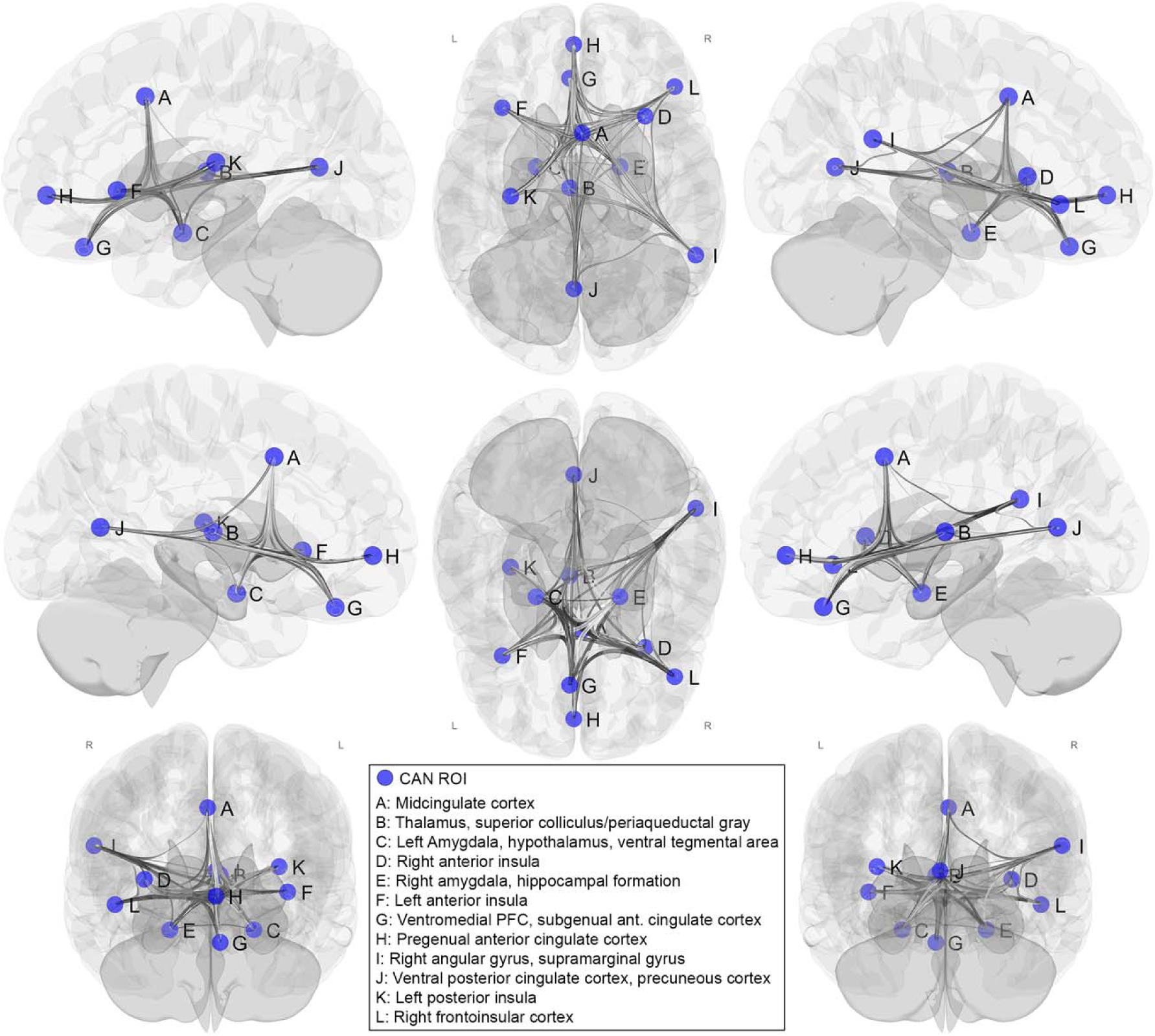
Resting state Central Autonomic Network (rsCAN) connectivity regions of interest (ROI). CAN ROIs are defined by 5mm spheres within the A: midcingulate cortex, B: thalamus, C: left amygdala/hypothalamus, D: right anterior insula, E: right amygdala/hippocampal formation, F: left anterior insula, G: ventromedial prefrontal cortex/subgenual anterior cingulate cortex, H: pregenual anterior cingulate cortex, I: right angular gyrus/supramarginal gyrus, J: ventral posterior cingulate cortex/precuneus/lingual gyrus, K: left posterior insula, and L: right frontoinsular cortex. ROI: region of interest, PFC: prefrontal cortex, CAN: central autonomic network.

**Table 1:** Montreal Neurological Institute coordinates defining central autonomic network (CAN) regions of interest centroids used in the present analysis.

| ROI | x | y | z |
| --- | --- | --- | --- |
| Midcingulate Cortex | 2 | 10 | 40 |
| Thalamus (medial-dorsal nucleus, pulvinar), Superior colliculus/periaqueductal gray | -4 | -16 | 8 |
| Left amygdala/hypothalamus | -20 | -6 | -18 |
| Right anterior Insula | 32 | 18 | 6 |
| Right amygdala/hippocampal formation | 20 | -6 | -18 |
| Left anterior Insula | -36 | 22 | 0 |
| Ventromedial PFC, subgenual ACC | -4 | 36 | -24 |
| Pregenuel ACC | -2 | 52 | -2 |
| Right angular gyrus, supramarginal gyrus | 56 | -48 | 22 |
| Ventral PCC, precuneous cortex, lingual gyrus | -2 | -64 | 10 |
| Left posterior Insula | -32 | -20 | 12 |
| Right frontoinsula cortex | 46 | 32 | -6 |

PFC: prefrontal cortex, ACC: Anterior cingulate cortex, PCC: posterior cingulate cortex

### fMRI rsCAN Connectivity Analysis

Functional and anatomical data were preprocessed using a flexible preprocessing pipeline in CONN (RRID:SCR_009550) release 22.a and SPM (RRID:SCR_007037) release 12.7771 (*49*), (**Supplementary Methods**). ROI-to-ROI connectivity matrices (RRC) were estimated characterizing the patterns of functional connectivity within 12 ROIs. Functional connectivity strength was represented by Fisher-transformed bivariate correlation coefficients from a weighted general linear model (weighted-GLM (*50*)), defined separately for each pair of target areas, then averaged. To compensate for possible transient magnetization effects at the beginning of each run, individual scans were weighted by a step function convolved with an SPM canonical hemodynamic response function and rectified.

### Data Analysis

A total of 70 participants characterized by demographics, *APOE* genotyping (*APOE4* carriers n= 37 and non-carriers n= 33), BP, BPV, resting state functional connectivity, VRF burden, and WML volumetric data were included in the analysis. Data was screened for outliers (3 SD from mean), and one WML volume outlier was identified and removed (>3.5 SD from mean). All models included age, average SBP, sex, and VRF burden as covariates. The WML volume variable was right skewed, so a square root transformation was used to reduce skewness. The square root of WML volume mm^3^ and the square root of IC volume mm^3^ were then converted to a white matter volume fraction to control for differences in IC volume, as previously described (*51*).

First, multiple linear regression analyses were conducted separately in *APOE4* carriers and non-carriers. For significant findings, a simple mediation analysis was performed to determine whether rsCAN functional connectivity (m) mediated the effect of BP VIM (x) on WML volume (y). This analysis was performed using model 4 in the Hayes’ PROCESS macro. Third, a simple moderation analysis was performed to determine whether rsCAN functional connectivity (w) moderated the effects of BP VIM (x) on WML volume (y). This analysis was performed using model 1 in the Hayes’ Process macro. All analyses were performed in R version 4.2.2.

All previous analyses were then repeated using a dichotomized CSVD variable as the dependent variable in logistic regression analyses in participants with CSVD characterization (*APOE4* carriers n=33 and non-carriers n= 26). Additionally, all analyses were repeated using BP SD and BP CV instead of BP VIM as independent variables. These analyses are detailed in **Supplementary File 1**.

## Results

A total of 69 participants with complete data were included after outlier removal, as described above. Comparison of *APOE4* carriers and non-carriers on all study variables is displayed in **Table 2**. There were no significant differences between *APOE4* carriers and non-carriers in age, average SBP, BPV, WML volume, rsCAN connectivity, sex, or VRF burden.

**Table 2:** Descriptive statistics by *APOE4* carrier status (*APOE4* carriers vs. non-carriers)

| Total Sample | *APOE4* carriers (n=37) | non-carriers (n=32) |  |
| --- | --- | --- | --- |
|  | Mean±SD | Mean±SD | *P* value |
| Age (range) | 69.81±6.96 (60-89) | 70.22±7.79 (55-88) | .82 |
| Average SBP | 136.60±15.17 | 139.08±23.06 | .61 |
| BP (VIM) | 5.3±0.7 | 5.3±0.8 | .90 |
| rsCAN Connectivity | .0489±.0283 | .0486±.0337 | .96 |
| WML/IC volume | 0.002 ± .0.0008 | 0.001±.0.0007 | .29 |
|  | Frequency (%) | Frequency (%) |  |
| Female | 25 (68) | 20 (63) | .85 |
| >1 VRF | 18 (49) | 17 (53) | .90 |
| Hypertension | 14 (38) | 14 (44) | .80 |
| Hyperlipidemia | 19 (51) | 17 (53) | .53 |
| Type II Diabetes | 5 (14) | 3 (9) | .87 |
| Smoking History | 16 (43) | 11 (34) | .61 |
| CVD | 4 (11) | 4 (13) | >.99<sup>a</sup> |
| Atrial fibrillation | 1 (3) | 2 (6) | .59<sup>a</sup> |
| TIA | 0 (0) | 0 (0) |  |
| CSVD Data Subset | *APOE4* carriers (n=33) | non-carriers (n=26) |  |
|  | Frequency (%) | Frequency (%) |  |
| Presence of CSVD<sup>b</sup> | 16 (48) | 14 (54) | .88 |
| Fazekas Score $\geq 2$ | 10 (30) | 5 (19) | .39 |
| Lacunes present | 4 (12) | 6 (23) | .55 |
| Microbleeds present | 6 (18) | 8 (31) | .45 |
| PVS<sup>c</sup> | 6 (18) | 8 (31) | .41 |

SBP: systolic blood pressure, WML: white matter lesion, IC: intracranial, BPV: blood pressure variability, VIM: variability independent of the mean, VRF: vascular risk factor, CVD: cardiovascular disease, TIA: transient ischemic attack, SD: standard deviation, PVS: perivascular spaces. P-values for continuous variables obtained using independent t-tests, p-values for count data obtained using chi-square tests. a: Fisher's exact test, b: Total CSVD score $\geq 1$ , c: moderate to severe basal ganglia perivascular space enlargement.

### Blood Pressure Variability (BPV) and CSVD Burden

Higher BP VIM was significantly associated with greater odds of CSVD presence and greater CSVD burden indexed by WML volume in *APOE4* carriers (*B*= 18.92, *P*= .02; *B*= .003, *P*= .04, respectively; **Table 3**), but not in non-carriers (*B*= 1.45, *P*= .83; *B*= -.001, *P*= .41, respectively) adjusted for age, average SBP, rsCAN connectivity, sex, and VRF. Analyses using BP SD and BP CV instead of BP VIM yielded similar results (**Supplementary File 1**).

**Table 3a:** Association between BP VIM and odds of CSVD feature presence in *APOE4* carriers adjusted for age, sex, average SBP, rsCAN connectivity, and VRF.

| Variable Name | *B* | standard error | *Z* | *P* value |
| --- | --- | --- | --- | --- |
| (intercept) | -26.36 | 10.69 | -2.47 | .01 |
| BP VIM | 18.92 | 8.28 | 2.28 | .02 |
| rsCAN | -22.66 | 19.64 | -1.15 | .25 |
| Average SBP | .06 | .04 | 1.49 | .14 |
| >1 VRF | 1.29 | 1.17 | 1.11 | .28 |
| Age | .13 | .08 | 1.50 | .13 |
| Sex (male) | -.52 | 1.09 | -.48 | .63 |

**Table 3b:** Association between BP VIM and WML volume in *APOE4* carriers adjusted for age, sex, average SBP, rsCAN connectivity, and VRF.

| Variable Name | *B* | standard error | *T* | *P value* |
| --- | --- | --- | --- | --- |
| (intercept) | -.006 | .001 | -4.09 | .0003 |
| BP VIM | .003 | .001 | 2.16 | .04 |
| rsCAN connectivity | -.002 | .004 | -.48 | .63 |
| Average SBP | .00001 | 0.000007 | 1.68 | .10 |
| VRF >1 | .0002 | .0002 | .80 | .43 |
| Age | .00006 | .00002 | 4.18 | .0002 |
| Sex (male) | -.0004 | .0002 | -1.56 | .13 |

**Dependent Variable 3a: CSVD feature presence, 3b: WML volume mm<sup>3</sup>/IC volume mm<sup>3</sup> fraction.**
BP: blood pressure, VIM: variability independent of the mean, SBP: systolic blood pressure, rsCAN: resting state central autonomic network, VRF: vascular risk factor, WML: white matter lesion, IC: intracranial.

### Mediation Analysis

Simple mediation analyses confirmed linear model findings that BP VIM was associated with odds of CSVD presence and CSVD burden indexed by WML volume in *APOE4* carriers adjusted for age, average SBP, VRF, sex and rsCAN connectivity (Figure 2). BP VIM was not associated with rsCAN functional connectivity in either the CSVD feature presence or WML volume analysis and rsCAN was not associated with CSVD feature presence or WML volume (Figure 2). The indirect effect was not significant in either analysis, indicating that the relationship between BPV and CSVD in *APOE4* carriers is direct and not mediated by rsCAN connectivity (Figure 2).

**Figure 2:**
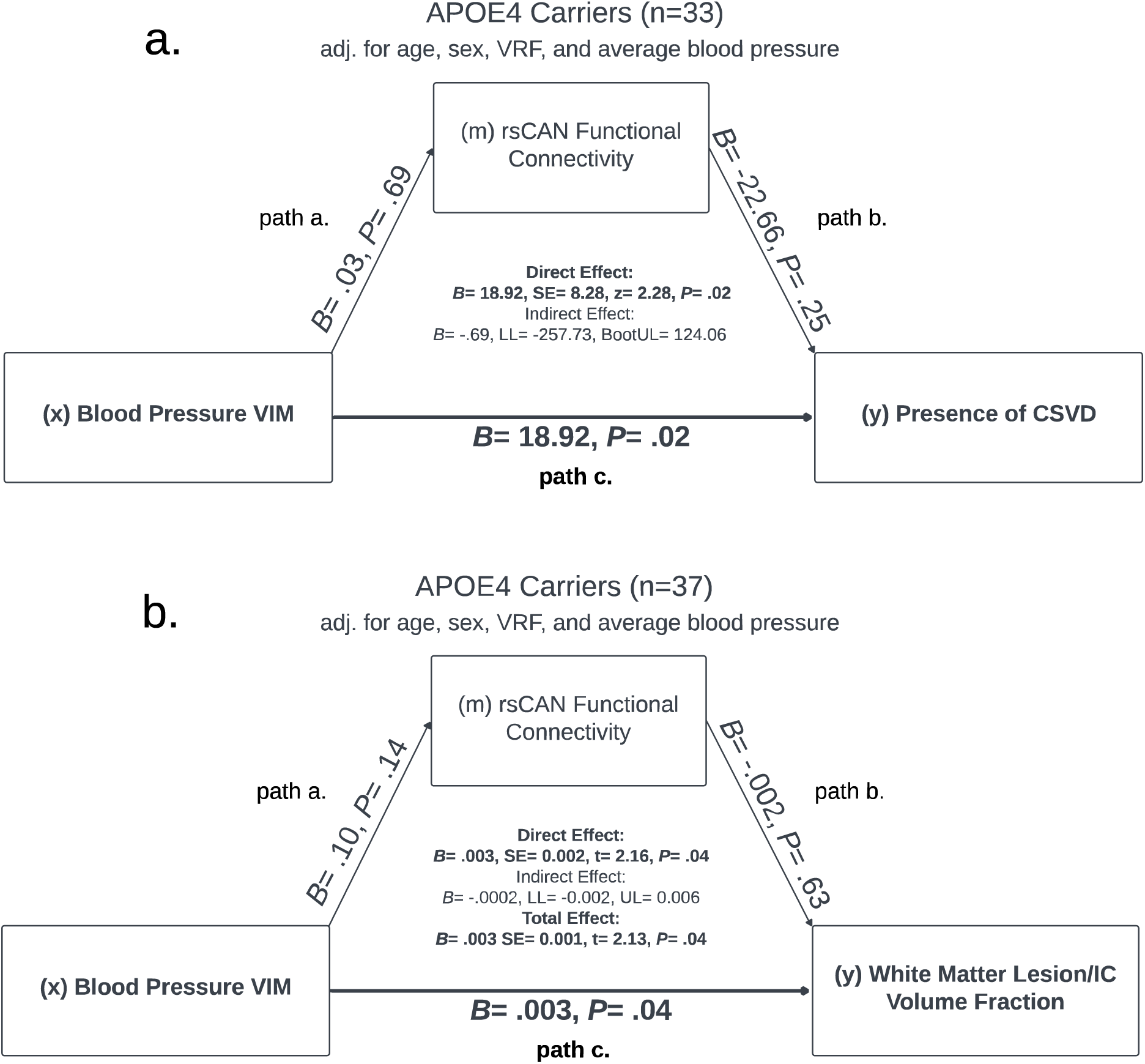
Panel a: Simple mediation analysis in 33 *APOE4* carriers. Unstandardized logistic regression coefficients (*B*) are displayed for the relationship between blood pressure variability independent of mean (VIM) and CSVD presence mediated by rsCAN functional connectivity. Path a= effect of VIM on rsCAN (linear regression coefficient), path b= effect of rsCAN on log odds of CSVD presence, path c= direct effect of VIM on log odds of CSVD presence adjusted for rsCAN, path a x b= indirect effect of VIM on log odds of CSVD presence. **Panel b:** Unstandardized linear regression coefficients (*B*) are displayed for the relationship between VIM and white matter lesion/intracranial (IC) volume fraction mediated by resting state central autonomic network (rsCAN) functional connectivity. All analyses are adjusted for age, sex, vascular risk factor burden (VRF), and average systolic blood pressure. Significant associations are indicated by bold font (p< .05). Both analyses performed using Hayes’ PROCESS macro model four. x= independent variable, y= dependent variable, m= mediator.

### Moderation Analysis

Simple moderation analysis confirmed BP VIM was associated with odds of CSVD presence and CSVD burden indexed by WML volume in *APOE4* carriers adjusted for age, average SBP, VRF, sex, and rsCAN connectivity (Figure 3). rsCAN connectivity was not significantly associated with WML volume or CSVD, however, there was evidence of an rsCAN*BPV interaction. Specifically, the relationship between higher BPV and greater WML volume/odds of CSVD was stronger in *APOE4* carriers with low rsCAN functional connectivity (Figure 3). The effect of BP VIM on odds of CSVD presence and WML volume in *APOE4* carriers at -1 SD, mean, and +1 SD rsCAN functional connectivity are shown in Figure 4.

**Figure 3:**
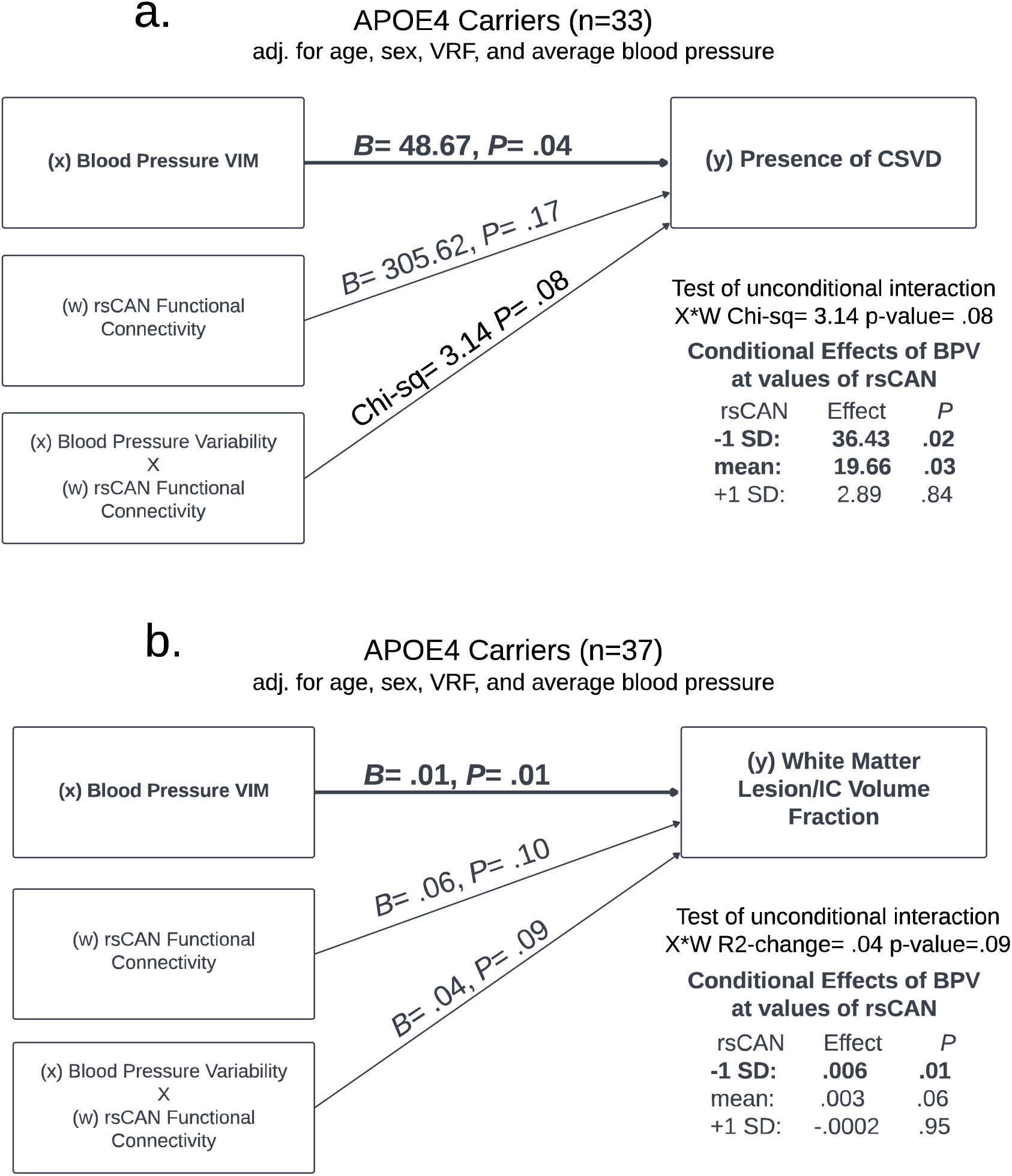
Panel a: Simple moderation analysis in 33 *APOE4* carriers showing unstandardized logistic regression coefficients (*B*) for the relationship between blood pressure variability independent of mean (VIM) and the presence of cerebral small vessel disease (CSVD), and the effect of interaction between rsCAN and VIM. **Panel b:** Simple moderation analysis in 37 *APOE4* carriers showing unstandardized linear regression coefficients (*B*) for the relationship between VIM and white matter lesion/intracranial (IC) volume fraction, and the effect of interaction between resting state central autonomic network connectivity (rsCAN) and blood pressure VIM. Conditional effects of VIM on WML volume and CSVD presence at -1 SD, mean, and +1 SD rsCAN are also displayed in each panel. Significant associations are indicated by bold font (*P*<.05) and all analyses are adjusted for age, vascular risk factor burden (VRF), average systolic blood pressure, and rsCAN functional connectivity. Both analyses performed using Hayes’ PROCESS macro model one. x= independent variable, y= dependent variable, w= moderator.

**Figure 4:**
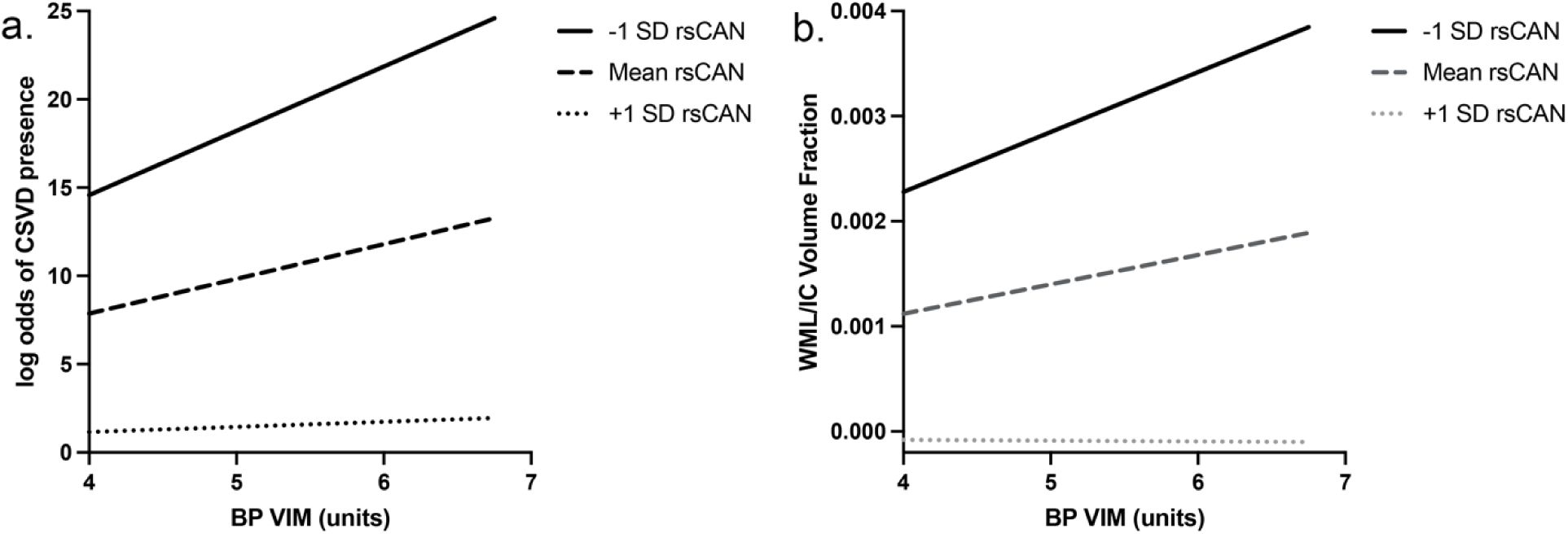
Panel a: Conditional effects plot showing the relationship between blood pressure (BP) variability independent of mean (VIM) and log odds of CSVD presence at three levels of resting state central autonomic network (rsCAN) connectivity: -1 SD, mean, and +1 SD. SD= standard deviation. **Panel b:** Conditional effects plot showing the relationship between BP VIM and white matter lesion (WML) volume percent of total intracranial volume at three levels of resting state central autonomic network (rsCAN) connectivity: -1 SD, mean, and +1 SD. SD= standard deviation. Both models adjusted for age, average SBP, sex, and VRF.

## Discussion

In the present study, older *APOE4* carriers with elevated beat-to-beat BPV exhibited increased CSVD presence and severity, independent of average blood pressure and CAN function. To our knowledge, this is the first study to report a relationship between BPV and CSVD, specifically in *APOE4* carriers. These findings are consistent with prior studies indicating stronger links between BPV and neuropathologic change in *APOE4* carriers versus non-carriers and underscores the vulnerability of *APOE4* carriers to vascular brain injury. Causal mediation analysis showed that changes in the CAN could not account for the relationship between BPV and CSVD presence or extent. Together these findings suggest that BPV is more likely a cause, rather than a consequence, of CSVD (*32, 33*).

To evaluate an alternative role for CAN dysfunction in exacerbating the adverse effects of BPV, rsCAN was also analyzed as a moderator of the relationship between BPV and WML volume. The relationship between higher BPV and greater CSVD burden was driven by *APOE4* carriers with lower rsCAN connectivity, implicating central autonomic function in BPV-associated risk for CSVD. Together these findings could suggest that BPV and CAN dysfunction may have a synergistic adverse effect on CSVD in older *APOE4* carriers (*9*). This result has clear implications for understanding BPV-related risk for vascular brain injury and supports further study of the interplay between CAN function and BPV in cerebrovascular disease.

Mounting research highlighting the role of BPV as a risk factor for cerebrovascular disease raises the question of what drives BPV elevation in older adults; however, the regulatory mechanisms underlying age-related changes in BPV are complex and involve baroreflex sensitivity (*52*), arterial stiffness (*53*), endothelial dysfunction (*54*), kidney function (*54*), and other factors. Adding complexity is the potential involvement of CAN regions, which themselves may be subject to damage from CSVD. For example, functional connectivity between the left amygdala, medial frontal gyrus, bilateral postcentral gyri, and bilateral paracentral lobules is thought to modulate baroreflex sensitivity (*27*), and baroreflex sensitivity is directly associated with BPV (*55, 56*). White matter damage to these network connections due to CSVD could disrupt CAN function, and by extension BPV regulation (*2*). However, our findings fail to support the hypothesis that BPV is a consequence of deteriorating CAN function. CAN functional connectivity was not associated with BPV in our analysis. If BPV was a consequence of CAN functional decline an inverse relationship might be expected instead. Also, CAN connectivity did not mediate the relationship between BPV and WML volume as might be expected if elevated BPV was associated with a decline in CAN function. Lastly, no significant difference in BPV was observed between *APOE4* carriers and non-carriers. Our findings instead reveal increased CSVD in *APOE4* carriers with BPV elevation and reduced CAN connectivity. *APOE4* carriers are also at increased risk for neurodegeneration, which may impact grey matter regions that underlie CAN functional connectivity. Future studies should similarly examine rsCAN mediation and moderation of the relationship between BPV and grey matter atrophy in *APOE4* carriers to gain additional insight into mechanisms underlying BPV-associated risk for dementia.

One of the strengths of the present study is the use of a CAN method derived from a voxel-based meta-analysis of 43 studies (*28*). Additional study strengths include use of multi- modal imaging to comprehensively characterize the presence or absence of CSVD by an aggregated analysis of WMLs, lacunes, cerebral microbleeds and enlarged perivascular spaces (*47, 57*), as well as objective quantification of the extent of CSVD burden by automated WML volumetrics. Examination of *APOE4* gene effects and mediation and moderation analyses are further strengths. Study limitations include the relatively small sample size and cross-sectional design, but this is the first study to test the hypothesis that CAN disruption may be involved in BPV effects on cerebrovascular disease, and the first to examine the relationship between BPV and CSVD in *APOE4* carriers and in non-carriers. Future longitudinal studies should be conducted to confirm these cross-sectional results.

The present study findings indicate CAN dysfunction may compound the adverse effect of BPV on CSVD in *APOE4* carriers, with substantial implications for our understanding of BPV- associated risk for cerebrovascular disease. Autonomic function is thought to play a role in cerebrovascular autoregulation, although there are many other contributing mechanisms (*58*). It is possible that autonomic dysfunction hampers the ability of the cerebrovasculature to adapt to large, rapid fluctuations in blood pressure (*58*), particularly in the context of genetic vulnerability to cerebrovascular disease (i.e., *APOE4*). Further study of specific autonomic and cerebrovascular autoregulatory functions related to low rsCAN connectivity, and BPV- associated risk for CSVD, may yield further insights with potential implications for treatment and prevention of CSVD in *APOE4* carriers at risk for dementia.

## Supporting information

Strobe Checklist

## Data Availability

The data that support the findings of this study are available upon reasonable request from the corresponding author, DN.

## Acknowledgements

This research was supported by National Institutes of Health grants (DAN: R01AG064228, R01AG060049, R01AG082073, P01AG052350), (SDH: K24AG081325) and the Canadian Institutes of Health Research (AK: DFD-170763).

