## Supplementary File 1 for "Blood pressure variability, central autonomic network dysfunction and cerebral small vessel disease in APOE4 carriers"

**Supplementary File 1: Supplementary results**

**Blood Pressure Variability and CSVD**

In logistic regression analyses, BPV (SD) was significantly associated with odds of CSVD presence in *APOE4* carriers (*B*= .48, *P*= .04; **Supplementary** **Table 1**), adjusted for age, average SBP, rsCAN connectivity, and VRF, but was not significantly associated with odds of CSVD presence in non-carriers (*B*= -.001, *P*= .99). Similar findings were observed in *APOE4 carriers* regardless of BPV metric.

**Supplementary Table 1:** Relationship between three different BPV metrics and CSVD presence adjusted for rsCAN, average SBP, VRF, and age in *APOE4* carriers.

| *BP Standard Deviation* | | | | |
| --- | --- | --- | --- | --- |
|  | *B* | std. error | z | *P* |
| (intercept) | -16.29 | 8.06 | -2.02 | .04 |
| BP SD | .48 | .23 | 2.11 | .04 |
| rsCAN | -21.09 | 19.33 | -1.09 | .28 |
| Avg SBP | .04 | .04 | 1.14 | .25 |
| >1 VRF | .98 | .98 | 1.00 | .32 |
| Age | .13 | .08 | 1.56 | .12 |
| *BP Coefficient of Variation* | | | | |
|  | *B* | std. error | z | *P* |
| (intercept) | -18.69 | 8.61 | -2.17 | .03 |
| BP CV | .68 | .32 | 2.12 | .03 |
| rsCAN | -20.75 | 19.07 | -1.09 | .28 |
| Avg SBP | .05 | .04 | 1.49 | .14 |
| >1 VRF | .94 | .98 | .96 | .34 |
| Age | .13 | .08 | 1.61 | .11 |

**Dependent variable: CSVD feature presence.**

BP : blood pressure, VIM: variability independent of the mean, SD: standard deviation, CV: coefficient of variability, rsCAN: resting state central autonomic network connectivity, avg SBP: average systolic blood pressure, VRF: vascular risk factor burden.

**Moderation Analysis**

Simple moderation analysis confirmed increased BP SD was associated with increased odds of CSVD presence in *APOE4* carriers (*B*= 2.37, *P*= .04). rsCAN connectivity was not significantly associated with CSVD presence (B= 132.83, *P*= .16), however, there was evidence of an rsCAN*BPV interaction. Similar findings were observed using BP CV as the measure of BPV. These results are shown in **Supplementary** **Figure 1**

**Supplementary Figure 1: Panel a:** Simple moderation analysis in 33 *APOE4* carriers showing unstandardized logistic regression coefficients (*B*) for the relationship between blood pressure standard deviation (BP SD) and the presence of cerebral small vessel disease (CSVD), and the effect of interaction between rsCAN and BP SD. **Panel b:** The same analysis was repeated using blood pressure coefficient of variability (CV). Conditional effects of BPV on odds of CSVD presence at -1 SD, mean, and +1 SD rsCAN are also displayed in each panel. Significant associations are indicated by bold font (*P*<.05) and all analyses are adjusted for age, vascular risk factor burden (VRF), and average systolic blood pressure. Both analyses performed using Hayes’ PROCESS macro model one. x= independent variable, y= dependent variable, w= moderator.

**BPV and WML Volume Fraction**

In a multiple linear regression analysis of all participants (n=69) BP SD was not associated with WML volume (*B*= 3.6e-05 *P*=.26) adjusted for age, average SBP, vascular risk factor burden and rsCAN connectivity. In a stratified analysis based on APOE4 carrier status (non-carriers n=32, carriers n= 37) BP SD was not significantly associated with WML volume in non-carriers (*B*= -4.4e-05 *P*= .35) and in carriers (*B*= 8.5e-05 *P*= .06).

**Supplementary Table 3: Association between blood pressure variability and white matter lesion volume in *APOE4* carriers adjusted for age, average systolic blood pressure, rsCAN connectivity, and vascular risk factor burden.**

| BP Standard Deviation | | | | |
| --- | --- | --- | --- | --- |
| Coefficients | *B* | standard error | t | *P value* |
| (intercept) | -4.7e-03 | 1.4e-03 | -3.31 | .0002 |
| BPV (SD) | 8.5e-05 | 4.4e-05 | 1.93 | .06 |
| Age | 6.8e-05 | 1.6e-05 | 4.34 | .0001 |
| Average SBP | 7.5e-06 | 7.1e-06 | 1.06 | .30 |
| rsCAN connectivity | -1.3e-03 | 3.9e-03 | -.33 | .74 |
| VRF >1 | 8.0e-05 | 2.1e-04 | .38 | .70 |
| BP Coefficient of Variation | | | | |
| Coefficients | *B* | standard error | t | *P value* |
| (intercept) | -5.0e-03 | 1.4e-03 | -3.55 | .001 |
| BPV (CV) | 1.2e-04 | 6.3e-05 | 1.84 | .08 |
| Age | 6.7e-05 | 1.6e-05 | 4.30 | .0002 |
| Average SBP | 1.0e-05 | 6.9e-06 | 1.49 | .15 |
| rsCAN connectivity | -1.3e-03 | 3.9e-03 | -.34 | .74 |
| VRF >1 | 6.2e-05 | 2.1e-04 | .30 | .77 |

**Dependent Variable: WML volume/IC volume fraction.**

BPV: blood pressure variability, SD: standard deviation, CV: coefficient of variation, SBP: systolic blood pressure, rsCAN: resting state central autonomic network, VRF: vascular risk factor, WML: white matter lesion, IC: intracranial.

**Moderation Analysis**

Simple moderation analysis showed increased BPV was associated with increased WML volume in *APOE4* carriers (*B*= .0002, *P*= .02). rsCAN connectivity was not significantly associated with WML volume, but there was evidence of BPV*rsCAN interaction, specifically, the relationship between higher BPV and greater WML volume was stronger in *APOE4* carriers with lower rsCAN functional connectivity (BP SD: -1 SD rsCAN: effect= .0002, *P*= .01, BP CV: -1 SD rsCAN: effect= .0002, *P*= .01). Similar results were seen across BPV measures.

**Supplementary Figure 2: Panel a:** Simple moderation analysis in 37 *APOE4* carriers showing unstandardized linear regression coefficients (*B*) for the relationship between blood pressure standard deviation (BP SD) and white matter lesion/intracranial (IC) volume ratio, and the effect of interaction between resting state central autonomic network connectivity (rsCAN) and BP SD. **Panel b:** The same analysis was then repeated using blood pressure coefficient of variability (CV). Conditional effects of BP variability on WML volume at -1 SD, mean, and +1 SD rsCAN are also displayed in each panel. Significant associations are indicated by bold font (*P*<.05) and all analyses are adjusted for age, vascular risk factor burden (VRF), and average systolic blood pressure. Both analyses performed using Hayes’ PROCESS macro model one. x= independent variable, y= dependent variable, w= moderator.
