## Supplementary File 2 for "Blood pressure variability, central autonomic network dysfunction and cerebral small vessel disease in APOE4 carriers"

**Supplementary File 2: fMRI preprocessing methods**

Functional and anatomical data were preprocessed using a flexible preprocessing pipeline in CONN (RRID:SCR_009550) release 22.a and SPM (RRID:SCR_007037) release 12.7771 (*1*), including realignment with correction of susceptibility distortion interactions, slice timing correction, outlier detection, direct segmentation and MNI-space normalization, and smoothing. Functional data were realigned using SPM realign & unwarp procedure (*2*), where all scans were co-registered to a reference image (first scan of the first session) using a least squares approach and a 6 parameter (rigid body) transformation, and resampled using b-spline interpolation to correct for motion and magnetic susceptibility interactions. Temporal misalignment between different slices of the functional data (acquired in interleaved Siemens order) was corrected following SPM slice-timing correction (STC) procedure (*3*), using sinc temporal interpolation to resample each slice BOLD timeseries to a common mid-acquisition time. Potential outlier scans were identified using ART as acquisitions with framewise displacement above 0.9 mm or global BOLD signal changes above 5 standard deviations (*4*), and a reference BOLD image was computed for each subject by averaging all scans excluding outliers. Functional and anatomical data were normalized into standard MNI space, segmented into grey matter, white matter, and CSF tissue classes, and resampled to 2 mm isotropic voxels following a direct normalization procedure (*5*) using SPM unified segmentation and normalization algorithm (*6*) with the default IXI-549 tissue probability map template. Last, functional data were smoothed using spatial convolution with a Gaussian kernel of 8 mm full width half maximum (FWHM).

In addition, functional data were denoised using a standard denoising pipeline including the regression of potential confounding effects characterized by white matter timeseries (5 CompCor noise components), CSF timeseries (5 CompCor noise components), motion parameters and their first order derivatives (12 factors) (*7*), outlier scans (below 75 factors) (*8*), session effects and their first order derivatives (2 factors), and linear trends (2 factors) within each functional run, followed by bandpass frequency filtering of the BOLD timeseries between 0.008 Hz and 0.09 Hz. CompCor (*9*) noise components within white matter and CSF were estimated by computing the average BOLD signal as well as the largest principal components orthogonal to the BOLD average, motion parameters, and outlier scans within each subject's eroded segmentation masks. From the number of noise terms included in this denoising strategy, the effective degrees of freedom of the BOLD signal after denoising were estimated to range from 19.2 to 163.3 (average 66.3) across all subjects.
